# The Multidimensional Assessment of Parenting Scale: Youth Report Form in a Clinical Sample

**DOI:** 10.1101/2024.10.02.24314779

**Authors:** Christina M Hogan, Emily Beckmann, Micaela Maron, Kelsey Sutton, April Highlander, Melissa Pielech, Jennifer C Wolff, Thamara Davis, Justin Parent

## Abstract

**Objective:** The Multidimensional Assessment of Parenting Scale (MAPS) was developed to assess a wide range of behaviors across positive and negative domains of parenting. This study aims to expand the utility of the MAPS by evaluating a youth-report version which provides an additional perspective on parenting practices.

**Method:** The study evaluated the youth-report form of the MAPS (MAPS-Y) in a large clinical population (*N* = 628) ranging from middle childhood (8-12) to adolescence (13-17) who were admitted to partial and inpatient psychiatric units. Youth and their caregivers completed the parent and youth versions of the MAPS questionnaire, and measures of child and adolescent psychopathology, emotion regulation, family context, and adversity. Analyses of factor structure, reliability, agreement, and validity were performed. The study also examined a short form of the MAPS-Y for reliability and validity.

**Results:** CFA and model fit indices indicated that all items loaded as expected onto subscales and with good fit. Analyses support strong reliability. The factor structure of the youth-report was invariant across developmental stages, included both positive and negative domains, and demonstrated strong psychometric properties. The MAPS-Y short form demonstrated strong validity and reliability.

**Conclusion:** The youth-report of the MAPS and its short form are appropriate for use among children and adolescents experiencing acute clinical symptoms. The MAPS youth-report will allow for nuanced, in-depth assessment of the parenting behaviors beyond parent-report that are critical to treatment outcomes in youth.

---

Parenting practices are primary targets of evidence-based psychosocial treatments for child and adolescent psychopathology, including conduct problems,^1–2^ autism,^3^ attention-deficit/hyperactivity disorder (ADHD),^4–6^ anxiety,^7–8^ obsessive-compulsive disorder (OCD),^9^ trauma,^10^ and depression.^11–12^ In addition to research identifying parenting as a critical mechanism of change in youth clinical outcomes, further evidence has demonstrated that parenting behaviors mediate the association between child symptoms and known risk factors such as parental symptomology. Parenting has been associated with children’s overall functioning, as well as disorders including ADHD, oppositional defiant disorder (ODD), anxiety, and depression, and across more broad symptoms of internalizing and externalizing.^13–16^

Despite ample evidence of parenting as a primary target in many youth psychosocial treatments and parenting practices as a clear mechanism of change in outcomes, formal assessment of parenting is often overlooked in clinical practice in favor of focusing on youth symptom outcomes.^17–19^ The gold standard in parenting assessment, observational methods, is difficult to implement in standard care due to limitations on time and physical location. Further, live observation in a clinical care context may not fully capture a naturalistic range of parenting behaviors (e.g., yelling, physical punishment) due to clinician presence, either in-person or on telehealth platforms. Therefore, the most common method used when assessing parenting is parent-reported surveys. However, these surveys have historically demonstrated weak psychometric properties (e.g., low reliability, limited variability, or ceiling effects).^17^ Parent-report surveys also require multiple scales to assess a range of parenting skills that are positive (e.g., warmth, supportiveness) and/or negative (e.g., hostility, laxness), and do not easily capture parenting across developmental stages.

The Multidimensional Assessment of Parenting Scale (MAPS) was developed to overcome these limitations of parent-report parenting scales.^20^ Using an empirically-based approach, the MAPS was created using items adapted from several well-established parenting scales. The MAPS factor structure includes a broadband positive parenting and a broadband negative parenting domain. Broadband positive parenting is comprised of four narrow-band scales: warmth, praise, proactive parenting, and supportiveness. Broadband negative parenting is comprised of three narrowband subscales: hostility, lax control, and physical control. The MAPS factor scores have shown strong psychometric properties, measurement invariance across three developmental stages (early childhood, middle childhood, and adolescence), and growing evidence for validity.^21^

Although the MAPS successfully addressed important limitations of prior parent-reported parenting scales by examining a wide range of behaviors across positive and negative domains, over-reliance on parent reports alone remains a significant limitation. Youth perceptions of parenting are a critical aspect of parenting assessment, given that observed parenting behavior has been shown to converge more with youth reports than with parent reports in the MAPS. For example, the correlation between observed positive parenting and youth-reported positive parenting is twice as large as the correlation with parent-report. Assessment of negative parenting has shown to be more nuanced, with the more “accurate” informant (i.e., higher correlation with observed negative parenting) depending on individual differences in parents and youths, such as experiences of psychopathology symptoms. For example, youth who report higher depressive symptoms may overreport negative parenting relative to observed levels, whereas the opposite is true for parenting reports. Ultimately, parenting assessment is likely incomplete without assessing perceptions from both parent and youth reports, but a full assessment of parenting practices that considers multiple informants has not been available. Thus, evidence-based assessment of parenting is a vital practice for monitoring progress and evaluating outcomes of family-based treatments.

The current study aimed to expand the utility of the MAPS by adapting and evaluating it for youth-report, which provides an additional perspective on parenting practices. We evaluated the youth-report form of the MAPS in a large clinical population ranging from middle childhood (8-12) to adolescence (13-17). We hypothesized that the MAPS-Y (youth) form would demonstrate a similar factor structure as the parent form (warmth, praise, proactive parenting, and supportiveness grouped in positive parenting; hostility, lax control, and physical control grouped in negative parenting). Further, we hypothesized similarly strong reliability and initial support for validity. Next, we explored the MAPS-Y’s measurement invariance across developmental stages and its sensitivity to family-based intervention. Finally, to address the limitation of time constraints in a clinical setting, we adapted and tested a short-form of the MAPS-Y.

## Method

### Participants

A total of 628 youth who were admitted to intensive psychiatric units were included in the current study. Units included a child (7-12 years old) partial hospitalization program (CPHP) and an adolescent (11-18 years old) inpatient unit (AIU). CPHP participants were admitted between September 2022 and September 2023, and AIU participants were admitted between October 2021 and June 2023. Overall, youth across both samples (*M* age = 14, *SD* = 2.56) self-identified as cisgender female (41.0%), cisgender male (31.5%), or transgender, non-binary, or otherwise gender non-conforming (27.4%). Youth sex assigned at birth as reported on their hospital medical records was 36% male and 64% female. Further, youth race was self-reported as American Indian or Indigenous (6.1%), Asian (3.3%), Black (17.4%), and White (63.1%), and youth ethnicity was self-reported as Latinx or Hispanic (29.5%) and non-Latinx or Hispanic (70.5%).

### Procedures

Participants completed the clinical assessment battery of measures within the first week of admission to one of the hospital programs. The hospital Institutional Review Board approved the study as a retrospective chart review, and the measures were administered to inform clinical care in inpatient and partial hospitalization programs. In the CPHP, parents also completed parallel measures of parenting and child functioning. If the participants had more than one hospitalization during the study time frame, only the initial admission was evaluated. Specific measures included in the chart review from both hospitalization programs are detailed below.

### Measures

#### Multidimensional Assessment of Parenting Scale – Parent and Youth Report Forms

(MAPS).^20^ The MAPS assesses positive and negative parenting practices across seven domains (34 total items). The scale was developed from established measures of parenting practices to select optimal parenting items constituting both positive and negative dimensions of warmth/hostility and behavioral control appropriate for parents of children across the developmental span from young childhood through adolescence. The MAPS has demonstrated excellent internal and test-retest reliability as well as strong support for the validity of MAPS subscale scores.^20,22–24^ Additionally, measurement invariance across youth developmental stages from young childhood to adolescence has been established for the MAPS subscales. The Youth-report form of the MAPS is a direct adaptation of the MAPS to orient the questions to the youth perspective on the same parenting questions.

The Broadband Positive Parenting subscale of the MAPS includes four narrowband subscales: Proactive Parenting which measures child-centered appropriate responses to anticipated difficulties; Positive Reinforcement which measures contingent responses to positive child behavior with praise, rewards, or displays of approval; Warmth which measures displays of affection; and Supportiveness which measures displayed interest in the child, encouragement of positive communication, and openness to a child’s ideas and opinions.

The Broadband Negative Parenting factor includes three narrowband subscales: Hostility which includes items representing intrusive parenting that is over controlling and parent-centered as well as harshness which includes coercive processes such as arguing, threats, yelling, ineffective discipline, and irritability; Physical Control which includes items representing physical discipline both generally and specifically out of anger and frustration; and Lax Control which includes items representing permissiveness or the absence of control, easily coerced control in which the parent backs down from control attempts based on the child’s behavior, and inconsistency which is the failure to follow through with control or inconsistent applying consequences.

### Adolescent (AIU) Validity Measures

#### Psychopathology Symptoms

The Suicidal Ideation Questionnaire-Junior (SIQ-JR)^25^ was used to measure past month suicidal ideation. Likert scale responses capture the frequency of thoughts and range from 0 (*I’ve never had this thought*) to 6 (*Almost every day*). The Self-Injurious Thoughts and Behaviors Interview (SITBI)^26^ was used to assess past month frequency of suicide attempts and non-suicidal self-injury. The Patient-Reported Outcomes Measurement Information System (PROMIS) short forms were used to assess anxiety, depression, and anger symptom severity.^27^ Each short form ranges from 4 to 8 items in length and asks respondents to assess the presence of their symptoms within the past 7 days. Scores for anxiety, depressive symptoms, and anger are calculated using the same 5-point rating scale ranging from “Never” to “Almost Always”.

#### Transdiagnostic Factors

The 18-item short form of the Difficulties in Emotion Regulation Scale (DERS)^28^ was used to assess domains of emotion regulation on a 1 (Almost never) to 5 (Almost always) Likert Scale. Responses summed for a total score, with higher scores indicating greater difficulties with emotion regulation. The short-form PROMIS Stress and Sleep Disturbances scales were used to assess subjective experiences of stress and difficulties with sleep quality or staying asleep. The PROMIS psychological stress^29^ and sleep disturbance^30^ scales use 5-point rating scales that range from 1 (Never) to 5 (Always). Higher scores indicate higher levels of stress or sleep disturbances.

#### Family Context and Adversity

The 12-item version of the Family Assessment Device (FAD-12)^31^ was used to assess adolescent satisfaction with general family functioning. A 4-point Likert scale rated responses from “Strongly Disagree” to “Strongly Agree”. Higher scores indicate more problematic overall family functioning. The Adverse Childhood Experiences Questionnaire (ACE-Q)^32^ was used to assess 19 adverse childhood experiences. Adolescents indicated how many items they have experienced, and a total count of items experienced was used in the current study.

### Child (CPHP) Validity Measures

#### Psychopathology Symptoms

The total score of the Screen for Child Anxiety Related Disorders (SCARED)^33^ was used to assess youth anxiety severity. A 3-point Likert scale ranged from “Not True or Hardly Ever True” to “Very True or Often True” with higher scores indicating more severe anxiety symptoms. The total score of the Children’s Depression Inventory 2nd Edition (CDI)^34^ was used to assess depressive symptom severity as well as a single item to assess suicidal ideation (“I do not think about killing myself”, “I think about killing myself but would not do it” or “I want to kill myself”). A 3-point item-specific Likert scale is used, with higher scores indicating more severe depressive symptoms. Parents completed the Disruptive Behavior Disorders (DBD)^35^ rating scale to assess externalizing symptoms. For the current study, the oppositional defiant disorder subscale was used to assess oppositionality, and an irritability scale was also created based on common irritability symptoms used in other scales (e.g., “Is often angry or resentful”). A 4-point Likert scale ranging from “Not at all” to “Very much” is used with higher scores indicating more severe oppositional or irritable symptoms.

#### Transdiagnostic Factors

Youth completed the Children’s Emotion Management Scale (CEMS-Youth Report).^36–37^ The emotional dysregulation subscales for worry, anger, and sadness were used to assess cross-cutting emotional dysregulation. Items are on a 3-point Likert-type scale ranging from “Hardly ever” to “Often” with higher scores indicating more severe emotion-specific dysregulation. Similar to the adolescent measures, the short-form of the PROMIS Sleep Disturbances scale was used to assess difficulties with sleep quality or staying asleep.^30^

#### Family Context and Adversity

The eight-item short-form of the PROMIS pediatric Family Relationships scale was used to assess perceptions of family connection and support. Items are rated on a 5-item Likert scale from “Never” to “Always” with higher scores reflecting higher quality family relationships. Parents completed the Center for Youth Wellness Adverse Childhood Experiences Questionnaire (CYW ACE-Q)^32,38^ to assess youth exposure to adversity.

A total of 17 ACEs were reported on, and a total score was used with higher scores indicating higher adversity.

### Data Analytic Plan

Analyses were conducted in four phases. In the first stage (model fit), confirmatory factor analysis was conducted based on the original parent-report factor structure of the MAPS. Analyses were conducted with the Lavaan R package using Jamovi statistical software. The following fit statistics were employed to evaluate model fit: chi-square, χ^2^ : p > .05 excellent, comparative fit index (CFI; > 0.90 acceptable, > 0.95 excellent), root mean square error of approximation (RMSEA; < 0.08 acceptable, < 0.05 excellent), and the standardized root mean square residual (SRMR; < 0.08 acceptable, < 0.05 excellent). We also aimed for factor loadings above .40. Following confirmatory factor analysis (CFA) models, we examined three forms of measurement invariance using multiple group CFAs: configural, metric, and scalar. Measurement invariance was examined across youth race, ethnicity, and developmental stage (i.e., 7-12 and 13-18).

In stage two (reliability and agreement), Omega and alpha coefficients were estimated for internal consistency. Further, for the CPHP youth sample only, we examined the cross-informant agreement using bivariate correlations between youth and parent-report forms (n = 99). In stage three (validity), we examined correlations between youth MAPS factor scores and psychopathology, transdiagnostic factors, and family context and adversity domains separately by CPHP (7-12) and AIU (11-18) samples, given that the measures used differed among hospital programs. Finally, in stage four (short-form), we developed a short form of the MAPS youth report using a combination of item response theory (IRT), CFA, correlations, and Omega reliability to ensure maximum coverage, reliability, and validity of a short-form. The CPHP and AIU samples were merged for analyses of primary model fit and reliability but examined separately for validity analyses.

## Results

### Phase One: Model Fit

The seven-factor structure mirroring the parent-report form demonstrated good model fit, χ^2^ (474) = 1339, *p* < .001, RMSEA = .054, 90% CI [.051, .057], CFI = .92, SRMR = .059, see Table 1 for complete CFA results. Furthermore, all items had a significant factor loading above .40 except for item 23 on the Lax Control subscale. Regarding measurement invariance, scale invariance was met for race and ethnicity (all χ^2^ difference test *p*s > .05) but not the developmental stage. For the developmental stage, only metric invariance was met (χ^2^ difference tests between metric and scale *p* < .05). However, ΔCFI was < .010, ΔRMSEA < .015, and ΔSRMR < .010 suggesting some support for strong invariance. Nevertheless, direct mean comparisons across developmental stages (e.g., t-tests) could introduce bias due to the lack of scalar invariance, and separate norms are warranted for middle childhood (7-12) and adolescence (13-17).

**Table 1.** Sociodemographic variables of study participants.

| **Variable** | **n** | **%** |
| --- | --- | --- |
| Child sex assigned at birth | Female | 64% |
|  | Male | 36% |
| Child gender | Cisgender female | 41.0% |
|  | Cisgender male | 31.5% |
|  | Transgender, non-binary, or otherwise gender non-conforming | 27.4% |
| Child race and Ethnicity | American Indian or Indigenous | 6.1% |
|  | Asian | 3.3% |
|  | Black | 17.4% |
|  | Latinx or Hispanic | 29.5% |
|  | White | 63.1% |

### Phase Two: Reliability and Agreement

Reliability was strong for all scales (see Table 2 for reliability, descriptive statistics, and correlations between subscales). Specifically, reliability was excellent for proactive parenting (Ω = .828), positive reinforcement (Ω = .847), warmth (Ω = .853), supportiveness (Ω = .831), hostility (Ω = .908), and physical control (Ω = .928) and was acceptable for lax control (Ω = .797). Reliability for the broadband positive (Ω = .925) and negative (Ω = .880) scales was also excellent. Regarding the cross-informant agreement, the correlation between youth and parent reports was modest for positive parenting (*r* = .220, *p* < .05) and negative parenting (*r* = .220, *p* < .05), which is consistent with past parent-youth agreement research on parenting^39–40^ and mental health.^41^

**Table 2.**
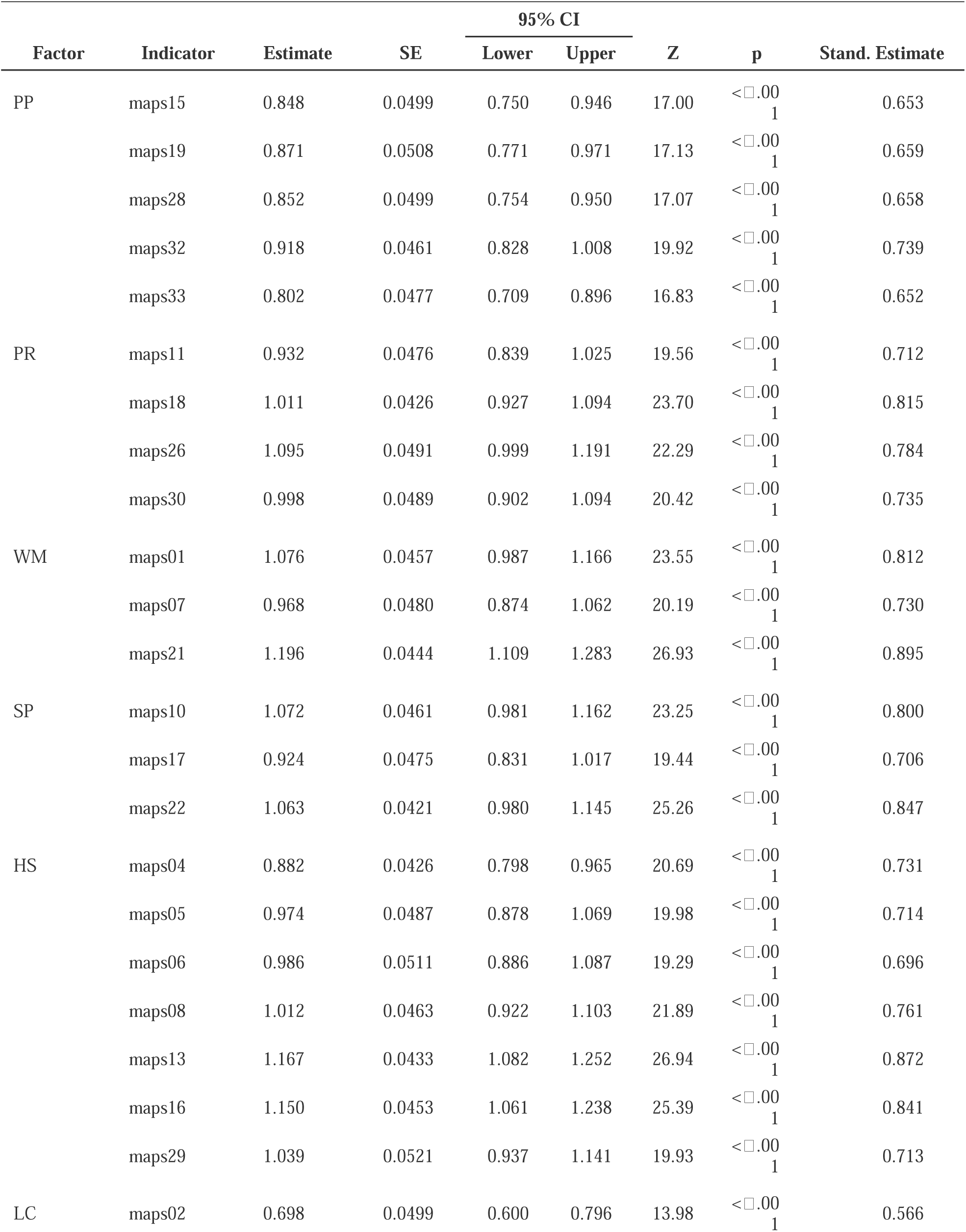
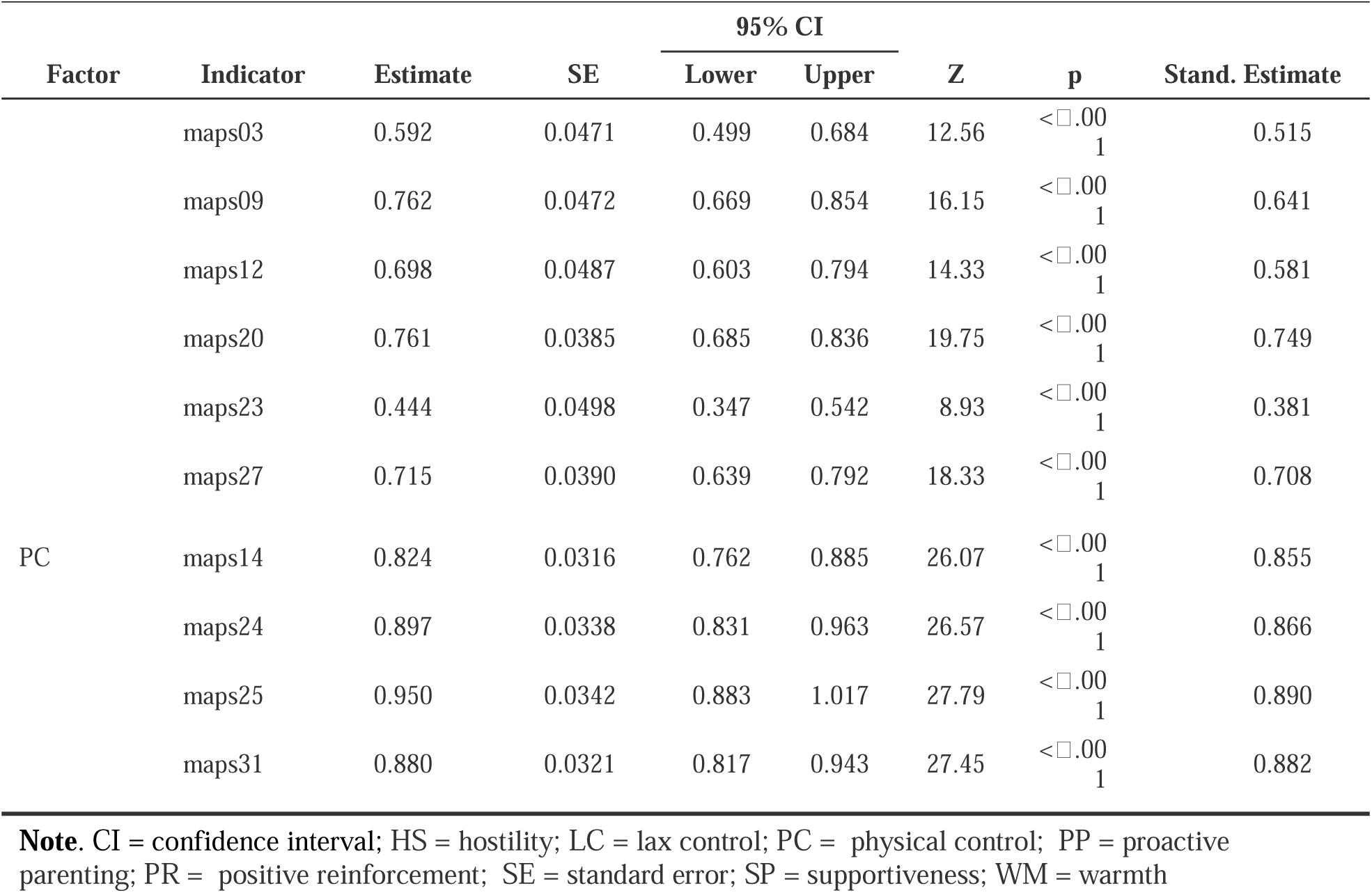
CFA item loadings for MAPS youth-form subscales.

### Phase Three: Validity

Tables 3 and 4 include correlations across the adolescent and child samples, respectively. In adolescence, higher levels of proactive parenting were associated with higher perceived stress, lower adversity, and lower family relationship quality. Unexpectedly, higher levels of proactive parenting were related to higher levels of past suicide attempts, though this could be consistent with a need for more proactive parenting practices when youth are at risk for self-harm. Higher levels of positive reinforcement, warmth, and supportiveness were related to higher adolescent self-compassion and perceived family relationship quality as well as lower levels of adversity. Further, higher levels of supportiveness were related to lower levels of adolescent emotion dysregulation. Regarding negative parenting, higher levels of hostility were associated with higher levels of depressive symptoms, suicidal ideation, suicide attempts, anxiety, irritability, emotion dysregulation, sleep disturbances, adversity, and stress. Further, higher hostility was also related to lower adolescent self-compassion and lower quality family relationships. More lax control was associated with higher levels of depressive symptoms, anxiety, irritability, and emotion dysregulation but was the only subscale not associated with family relationship quality or adversity. Finally, more physical control was associated with higher depressive symptoms, anxiety, irritability, emotion dysregulation, adversity, and sleep disturbance as well as lower levels of family relationship quality.

**Table 3.** Mean, standard deviations, reliability, and correlations between youth-report narrowband and broadband scales.

|  | M (SD) | Omega | PP | PR | WM | SP | HS | LC | PC | POS |
| --- | --- | --- | --- | --- | --- | --- | --- | --- | --- | --- |
| PP | 2.94 (.949) | .828 | — |  |  |  |  |  |  |  |
| PR | 3.07 (1.10) | .847 | 0.641 | — |  |  |  |  |  |  |
| WM | 3.10 (1.17) | .853 | 0.453 | 0.594 | — |  |  |  |  |  |
| SP | 3.22 (1.12) | .831 | 0.623 | 0.727 | 0.642 | — |  |  |  |  |
| HS | 2.82 (1.09) | .908 | -0.090 | -0.229 | -0.246 | -0.376 | — |  |  |  |
| LC | 2.04 (.753) | .797 | 0.247 | 0.268 | 0.174 | 0.151 | 0.213 | — |  |  |
| PC | 1.52 (.922) | .928 | -0.064 | -0.145 | -0.112 | -0.212 | 0.462 | 0.179 | — |  |
| POS | 3.06 (.892) | .925 | 0.788 | 0.879 | 0.815 | 0.891 | -0.285 | 0.250 | -0.161 | — |
| NEG | 2.23 (.684) | .880 | 0.014 | -0.089 | -0.115 | -0.242 | 0.824 | 0.567 | 0.767 | -0.127 |

**Note.** HS = hostility; LC = lax control; M = mean; NEG = broadband negative parenting; PC = physical control; POS = broadband positive parenting; PP = proactive parenting; PR = positive reinforcement; SD = standard deviation; SP = supportiveness; WM = warmth.

**Table 4a.** Adolescent inpatient unit validation: ages 13-18 years.

|  | PP | PR | WM | SP | HS | HSs | LC | LCs | PC | POS | POSs | NEG |
| --- | --- | --- | --- | --- | --- | --- | --- | --- | --- | --- | --- | --- |
| Depression | 0.077 | -0.033 | -0.015 | -0.082 | **0.349\*** | **0.301\*** | **0.107\*** | 0.084 | **0.116\*** | -0.020 | -0.048 | **0.279\*** |
| SITBI | **0.162\*** | 0.092 | 0.096 | 0.100 | **0.178\*** | **0.152\*** | 0.042 | -0.006 | -0.010 | **0.135\*** | 0.117 | **0.112\*** |
| SIQ | 0.057 | -0.063 | -0.021 | -0.079 | **0.309\*** | **0.249\*** | 0.065 | 0.021 | 0.067 | -0.032 | -0.051 | **0.222\*** |
| Anxiety | 0.081 | -0.018 | 0.011 | -0.070 | **0.321\*** | **0.280\*** | **0.108\*** | **0.091\*** | **0.162\*** | -0.002 | -0.024 | **0.285\*** |
| Irritability | 0.062 | 0.005 | -0.044 | -0.080 | **0.342\*** | **0.323\*** | **0.189\*** | **0.166\*** | **0.159\*** | -0.026 | -0.022 | **0.325\*** |
| DERS | 0.040 | -0.062 | -0.069 | **-0.143\*** | **0.371\*** | **0.326\*** | **0.159\*** | **0.127\*** | **0.131\*** | -0.075 | -0.080 | **0.315\*** |
| Self-com | 0.086 | **0.161\*** | **0.100\*** | **0.159\*** | **-0.195\*** | **-0.141\*** | 0.049 | 0.069 | **-0.099\*** | **0.156\*** | **0.144\*** | **-0.133\*** |
| Sleep | 0.016 | -0.045 | -0.059 | **-0.106\*** | **0.307\*** | **0.298\*** | 0.049 | 0.019 | **0.134\*** | -0.063 | -0.062 | **0.242\*** |
| Stress | **0.093\*** | -0.004 | 0.022 | -0.025 | **0.340\*** | **0.299\*** | **0.102\*** | 0.076 | 0.053 | 0.023 | -0.002 | **0.246\*** |
| ACE | **-0.127\*** | **-0.140\*** | **-0.220\*** | **-0.235\*** | **0.326\*** | **0.297\*** | -0.010 | -0.021 | **0.194\*** | **-0.215\*** | **-0.226\*** | **0.257\*** |
| FAD | **-0.387\*** | **-0.513\*** | **-0.540\*** | **-0.643\*** | **0.594\*** | **0.561\*** | -0.054 | -0.078 | **0.337\*** | **-0.614\*** | **-0.644\*** | **0.449\*** |

**Note.** ACE = adverse childhood experiences; DERS = Difficulties in Emotion Regulation Scale; FAD = family assessment device; HS = hostility; LC = lax control; NEG = broadband negative parenting; PC = physical control; POS = broadband positive parenting; POSs = POS short form; PP = proactive parenting; PR = positive reinforcement; SITBI = self-injurious thoughts and behaviors interview; Self-com = self-compassion; SP = supportiveness; WM = warmth

**Table 4b.** Child partial hospital program validation: ages 7-12 years.

|  | PP | PR | WM | SP | HS | HSs | LC | LCs | PC | POS | POSs | NEG |
| --- | --- | --- | --- | --- | --- | --- | --- | --- | --- | --- | --- | --- |
| Depression | **-0.211\*** | **-0.353\*** | **-0.351\*** | **-0.307\*** | **0.394\*** | **0.327\*** | 0.014 | -0.031 | **0.190\*** | **-0.388\*** | -0.380* | **0.289\*** |
| SI | **-0.236\*** | **-0.292\*** | **-0.298\*** | **-0.281\*** | **0.421\*** | **0.350\*** | 0.008 | -0.048 | **0.282\*** | **-0.348\*** | **-0.334\*** | **0.343\*** |
| Anxiety | **-0.059** | **-0.144\*** | **-0.289\*** | **-0.186\*** | **0.402\*** | **0.343\*** | **0.219\*** | 0.121 | **0.251\*** | **-0.217\*** | **-0.183\*** | **0.399\*** |
| Irritability | 0.008 | -0.065 | -0.117 | -0.110 | **0.202\*** | **0.182\*** | 0.156 | 0.117 | 0.091 | -0.092 | -0.056 | **0.197\*** |
| Oppositional | 0.031 | -0.089 | -0.136 | -0.130 | **0.252\*** | **0.243\*** | **0.218\*** | 0.19 | 0.108 | -0.105 | -0.058 | **0.252\*** |
| Worry Dysreg | 0.081 | 0.074 | 0.017 | 0.051 | 0.132 | 0.117 | 0.086 | 0.098 | 0.037 | 0.069 | 0.054 | 0.115 |
| Sad Dysreg | **0.174\*** | 0.003 | 0.162 | 0.041 | 0.140 | 0.097 | 0.123 | 0.082 | 0.071 | 0.111 | 0.094 | 0.149 |
| Anger Dysreg | -0.023 | -0.039 | -0.087 | -0.072 | **0.312\*** | **0.308\*** | 0.057 | 0.06 | **0.265\*** | -0.070 | -0.085 | **0.300\*** |
| Sleep | -0.113 | -0.108 | **-0.198\*** | -0.115 | **0.248\*** | **0.215\*** | 0.099 | 0.056 | 0.012 | **-0.167\*** | **-0.159\*** | **0.165\*** |
| ACE | 0.031 | -0.009 | -0.02 | 0.115 | -0.110 | -0.149 | 0.033 | -0.026 | -0.092 | 0.037 | 0.000 | -0.079 |
| Family | **0.300\*** | **0.552\*** | **0.515\*** | **0.570\*** | **-0.443\*** | **-0.382\*** | 0.072 | 0.109 | **-0.190\*** | **0.620\*** | **0.583\*** | **-0.281\*** |

**Note.** ACE = adverse childhood experiences; HS = hostility; LC = lax control; NEG = broadband negative parenting; PC = physical control; POS = broadband positive parenting; POSs = POS short-form; PP = proactive parenting; PR = positive reinforcement; SI = suicidal ideation; SP = supportiveness; WM = warmth

In childhood, higher levels of proactive parenting, positive reinforcement, warmth, and supportiveness were related to lower depressive symptoms, suicidal ideation, and anxiety as well as higher family relationship quality. In addition, higher levels of warmth were related to lower levels of sleep disturbances, and higher levels of proactive parenting were related to high sadness dysregulation. Similar to adolescent results, higher levels of hostility were related to higher child depressive symptoms, suicidal ideation, anxiety, irritability, and sleep disturbances as well as lower family relationship quality. Further, higher hostility was associated with higher oppositionality and anger dysregulation. Higher lax control was only associated with higher anxiety and oppositionality. Lastly, more physical control was related to higher depressive symptoms, suicidal ideation, anxiety, and anger dysregulation as well as lower family relationship quality.

### Phase Four: Short-Form Development of the MAPS-Y

We began development of the short-form by using an IRT graded response model for each narrowband subscale. We then selected items with the highest discrimination parameters for inclusion in the short-form. For positive parenting, we sought to create a single broadband scale given that narrowband scales were already short. Based on discrimination parameters, we selected six items for the broadband positive parenting short-form that covered each of the narrowband domains. Regarding negative parenting, we developed a short-form of the hostility and lax control subscales, each with three items with the highest IRT discrimination parameters. A total of 12 items were selected for the short-form. We then conducted a CFA with all three short-form scales and model fit was good, χ^2^ (51) = 224, *p < .*001, RMSEA = .074, 90% CI [.064, .083], CFI = .948, SRMR = .051. Across scales, standardized factor loadings ranged from .558 to .888 (see Table 5). Reliability was excellent for short-form positive parenting (Ω = .871) and hostility (Ω = .874) and was acceptable for lax control (Ω = .754). Correlations between full and short-form scales were *r* = .940 for positive parenting, *r* = .934 for hostility, and *r* = .894 for lax control. Parent-youth agreement for the short-form was similar and ranged from *r* = .256 to *r* = .317. Finally, patterns of correlations with youth and family outcomes were similar to the full version and are presented in Tables 3 and 4.

**Table 5.** Short-form confirmatory factor analysis results.

| Short-form Factor Loadings |  |  |  |  |  |  |  |  |
| --- | --- | --- | --- | --- | --- | --- | --- | --- |
| Factor | Indicator | Estimate | SE | 95% Confidence Interval |  | Z | p | Stand. Estimate |
|  |  |  |  | Lower | Upper |  |  |  |
| PosPar | maps22 | 1.027 | 0.043<sub>3</sub> | 0.942 | 1.111 | 23.7 | <□.00<sub>1</sub> | 0.819 |
|  | maps32 | 0.739 | 0.047<sub>8</sub> | 0.645 | 0.833 | 15.4 | <□.00<sub>1</sub> | 0.596 |
|  | maps18 | 0.932 | 0.044<sub>8</sub> | 0.844 | 1.020 | 20.8 | <□.00<sub>1</sub> | 0.751 |
|  | maps21 | 0.930 | 0.049<sub>0</sub> | 0.834 | 1.026 | 19.0 | <□.00<sub>1</sub> | 0.698 |
|  | maps10 | 1.043 | 0.047<sub>2</sub> | 0.951 | 1.135 | 22.1 | <□.00<sub>1</sub> | 0.779 |
|  | maps26 | 1.020 | 0.050<sub>9</sub> | 0.921 | 1.120 | 20.0 | <□.00<sub>1</sub> | 0.731 |
| HS | maps08 | 1.005 | 0.047<sub>4</sub> | 0.913 | 1.098 | 21.2 | <□.00<sub>1</sub> | 0.756 |
|  | maps13 | 1.188 | 0.045<sub>2</sub> | 1.100 | 1.277 | 26.3 | <□.00<sub>1</sub> | 0.888 |
|  | maps16 | 1.169 | 0.047<sub>0</sub> | 1.077 | 1.261 | 24.9 | <□.00<sub>1</sub> | 0.855 |
| LC | maps09 | 0.664 | 0.050<sub>2</sub> | 0.566 | 0.762 | 13.2 | <□.00<sub>1</sub> | 0.558 |
|  | maps20 | 0.789 | 0.042<sub>7</sub> | 0.705 | 0.872 | 18.5 | <□.00<sub>1</sub> | 0.776 |
|  | maps27 | 0.786 | 0.042<sub>3</sub> | 0.703 | 0.869 | 18.6 | <□.00<sub>1</sub> | 0.777 |

**Note.** HS = hostility; LC = lax control; PosPar = positive parenting; SE = standard error;

## Discussion

The current study evaluated the youth-report version of the MAPS with the overall goal of identifying a reliable and valid measure of parenting behavior as reported by a clinical sample of youths. Confirmatory factor analyses and model fit indices demonstrated that all items included in the survey loaded onto subscales as expected and with good fit, providing support for use of the same factor structure in the MAPS youth-report as in the parent-report version. As hypothesized, analyses demonstrated strong reliability and validity of the measure, establishing the youth-report version as a useful measure of parenting behaviors in a clinical sample.

An integral objective for the MAPS youth report was addressing the field’s current dearth of multi-informant measures of parenting behaviors. Analyses of convergence across reporters demonstrated modest agreement between parent and youth report for positive and negative parenting behaviors. These findings provide further confidence in the measure’s ability to address the scarcity of parenting assessments that collect the perspectives of both youth and parents. These results also align with existing research and theory for youth and parent agreement on parenting measures.^39,40^ In addition, the factor structure did not vary depending upon the age of the respondents (i.e., child or adolescent sample), indicating a significant strength of this measure as applicable across developmental stages.

Validity of the measure was tested by examining associations between MAPS youth-report subscales and youth-reported internalizing and externalizing symptoms. Contrary to hypotheses, not all subscales demonstrated strong associations with the expected symptoms across the child and adolescent samples. For example, in the adolescent sample, high hostility was related to high self-compassion and higher quality family relationships. In the child sample, high proactive parenting was related to high sadness dysregulation. Further, several associations were found between internalizing and externalizing symptoms in both adolescent and child samples. Among adolescents, high hostility on the negative parenting scale was related to higher anxiety and depression. Similarly, for the child sample, proactive parenting was associated with lower depression and anxiety. Additionally, high hostility was related to higher depression and anxiety. These correlations suggest that some symptoms may be more predictive of certain subscales. However, consistent associations were found between symptoms and both the hostility and broadband negative parenting scales, suggesting that parenting-based interventions may specifically target hostility within negative parenting. Finally, examination of a short form of the MAPS-Y demonstrated that the short form reliably measures each of the three parenting domains, shows agreement with parents and youths, and correlates with clinical measures comparably to the full version. These findings strengthen the MAPS-Y’s applicability and utility in clinical contexts, where time is often a constraint for assessments.

Limitations of the current study should also be noted. The use of a clinical, hospital-based sample allows for the much-needed use of the MAPS youth-report across acute and clinical populations, but the use of the measure in non-clinical samples is therefore not generalizable from these results. Future research should examine the MAPS youth-report in community samples to allow for the widespread use of a multi-informant parent behavior measurement across care settings. Additionally, this study combined responses from participants across childhood and adolescence. While this approach allows for broad application of the measure across developmental stages, future research may explore the factor structure individually in childhood and adolescence to determine the need for higher specificity of reporters (i.e., child version and adolescent version). Thus, another future direction may include assessing the measure’s sensitivity to change over development.

Taking these findings together, the current study demonstrates the youth-report and youth-report short form of the MAPS as appropriate for use among children and adolescents experiencing acute clinical symptoms. The factor structure of the youth-report was invariant across developmental stages, included both positive and negative domains, and evidenced strong psychometric properties. The development of the youth-report is a much-needed addition to the parent-reported MAPS, and its use will allow for nuanced, in-depth assessment of the parenting behaviors that are so critical to youth outcomes.

## Data Availability

Summary statistical information is available, but raw data is not available for sharing due to the chart review nature of the study.

## Acknowledgements

The last author was supported by NICHD (L40HD103048), and by the Bradley Hospital COBRE Center for Sleep and Circadian Rhythms in Child and Adolescent Mental Health (P20GM139743).

